# Severity of thermal burn injury is associated with systemic neutrophil activation

**DOI:** 10.1101/2021.10.07.21264679

**Authors:** Maria Laggner, Marie-Therese Lingitz, Dragan Copic, Martin Direder, Katharina Klas, Daniel Bormann, Alfred Gugerell, Bernhard Moser, Christine Radtke, Stefan Hacker, Michael Mildner, Hendrik Jan Ankersmit, Thomas Haider

## Abstract

**Objectives:** Burn injuries elicit a unique and dynamic stress response which can lead to burn injury progression. Though neutrophils represent crucial players in the burn-induced immunological events, the dynamic secretion pattern and systemic levels of neutrophil-derived factors have not been investigated in detail so far.

**Methods:** Serum levels of neutrophil elastase (NE), myeloperoxidase (MPO), citrullinated histone H3 (CitH3), and complement factor C3a were quantified in burn victims over 4 weeks post injury. Furthermore, the potential association with mortality, degree of burn injury, and inhalation trauma was evaluated. In addition, leukocyte, platelet, neutrophil, and lymphocyte counts were assessed. Lastly, we analyzed the association of neutrophil-derived factors with clinical severity scoring systems.

**Results:** Serum levels of NE, MPO, CitH3, and C3a were remarkably elevated in burn victims compared to healthy controls. Leukocyte and neutrophil counts were significantly increased on admission day and day 1, while relative lymphocytes were decreased in the first 7 days post burn trauma. Though neutrophil-derived factors did not predict mortality, patients suffering from 3^rd^ degree burn injuries displayed increased CitH3 and NE levels. Accordingly, CitH3 and NE were elevated in cases with higher abbreviated burn severity indices (ABSI).

**Conclusions:** Taken together, our data suggest a role for neutrophil activation and NETosis in burn injuries and burn injury progression. Targeting exacerbated neutrophil activation might represent a new therapeutic option for severe cases of burn injury.

## Introduction

Burn injury is an umbrella term for a trauma, most commonly affecting the skin or lung, caused by a variety of external challenges, such as thermal extremes, deleterious radiation, alkaline and acidic chemicals, or excessive friction (1). The respective cause and burn degree further dictate clinical management, such as surgical intervention and moist rewarming following heat- and cold-induced injuries, respectively. In spite of the development of new therapeutic approaches, burn injuries are still associated with a high mortality rate, most commonly resulting from multiple organ failure, sepsis, and respiratory complications (2). Morbidity and mortality can be increased up to 10 years after the initial insult (3).

Burn trauma elicits a unique systemic stress response characterized by increased metabolism and inflammation (4). The early systemic inflammatory response syndrome (SIRS) is characterized by pro-inflammatory cytokines, such as interleukin 6 (IL-6), IL-8, and tumor necrosis factor alpha (TNFα) and usually lasts for several days (5, 6). Later, the milieu is shifted towards an anti-inflammatory response syndrome (AIRS), where the immunosuppressant mediators TGFβ1, and IL-10 prevail (6). Though inflammation is inherent and indispensable for normal wound healing (7), burn injuries can induce a state of distorted inflammatory response which can persist up to several years (8) and which can ultimately lead to host tissue damage and organ dysfunction. Various factors orchestrate early immune reactions, such as the amount of affected body surface area, burn depth and cause, inhalation injury, patient age, and chronic medical conditions (9). Neutrophils (10) and macrophages (11) are early key players infiltrating the burn-injured area. Though these immune cells are readily activated by burn injury-induced damage-associated molecular patterns (DAMPs), macrophage antigen presentation and neutrophil-mediated killing of pathogens have been shown to be diminished following burn injury (12, 13), leading to increased susceptibility to infections. While immunological hyper-activation entails tissue damage, exaggerated immunosuppression predisposes to infections (14). Hence, a delicate balance of immunomodulators determines clinical outcome in burn injuries.

A commonly observed phenomenon of burn victims is secondary burn progression (15). While tissue necrosis and ischemia are immediate results of burn injury, tissue damage is often aggravated long after the initial trauma. Formation of red blood cell aggregates and microthrombi and subsequent occlusion of the microvasculature have been identified as the underlying cause of burn progression (16). Though secondary burn progression substantially promotes tissue damage, its role in the post injury tissue response is often underestimated.

Neutrophils are implicated in the host defense against pathogens, whereby antimicrobial activity is exerted by phagocytosis, release of effector molecules, and formation of neutrophil extracellular traps (NETs) (17). Activated protein-arginine deiminase 4 (PAD4) catalyzes citrullination of histones, which results in chromatin decondensation. Granule proteins, such as myeloperoxidase (MPO) and neutrophil elastase (NE), further promote DNA de-compaction and intracellular DNA together with granule proteins are released following plasma membrane rupture (18, 19). It was further demonstrated that neutrophils contain intracellular stores of C3a (20), which might be self-synthesized or absorbed from the serum (21). Moreover, C3a was reported to induce neutrophil activation (22). A role for neutrophils in coagulation and microvascular obstruction has been described at several occasions (23-27). The amount of neutrophil-derived circulating, free DNA was proposed as a predictor of mortality in severely burnt patients (28) and elevated human neutrophil elastase DNA and nucleosomes were detected in burn and sepsis patients (29). Though previous studies have reported a role for neutrophils and NETs in burn injury (30, 31), systemic surrogate markers indicating NETosis have not been comprehensively studied so far. Our group was previously able to show activation of the soluble suppressor of tumorigenicity 2 (sST2) / IL-33 axis in sepsis (32) as well as thermal burn injury (33) and found increased soluble ST2 to be a predictor of mortality. In the current study, we aimed to delineate the dynamics of neutrophil-derived immunomodulators to deepen our understanding of NETosis in the post-burn injury immune response.

## Materials and methods

### Ethical statements

This study was approved by the institutional review board of the Medical University of Vienna (Vienna, Austria) (vote 593/2011) and was conducted in accordance with the Declaration of Helsinki and applicable local regulations. Written informed consent was obtained from all donors.

### Patient cohort and serum sample acquisition

Samples used in the current study have already been analyzed for sST2 and IL-33 (33). Patients > 18 years who were admitted within 24 hours post trauma to burn intensive care unit and presented with a burn injury covering > 10 % of the total body surface area (TBSA) at primary survey were included in this study (Table 1). Patients with chronic infectious diseases or autoimmune disorders were excluded. Eight healthy volunteers served as controls.

**Table 1.** Study group demographics. ABSI abbreviated burn severity index, APACHE II acute physiology and chronic health evaluation II, F:M female to male ratio, LOH length of hospitalization, LOS length of stay at intensive care unit, SOFA sequential organ failure assessment, TBSA total body surface area.

|  | burn patients | controls |
| --- | --- | --- |
| n | 32 | 8 |
| age in years * | 51.9 (46.5) ± 21.9 [33 – 74] | 40.5 (36) ± 19.9 [23 – 56] |
| F:M ratio [%] | 10:22 (31.3:68.7) | 3:5 (37.5:62.5) |
| TBSA [%] * | 32.5 (30.0) ± 19.6 [16.3 – 39.5] | |
| LOH [days] * | 37.2 (27.5) ± 33.9 [10.5 – 61.5] | |
| LOS [days] * | 29.7 (19.5) ± 33.0 [7.3 – 35.3] | |
| SOFA * | 6.8 (6.5) ± 3.6 [5 – 9] | |
| APACHE II * | 19.7 (18.0) ± 9.3 [13.5 – 28.0] | |
| ABSI | 7.91 (8.0) ± 2.8 [5 – 9] | |
| deceased [%] § | 6 (18.8) |  |
| 3<sup>rd</sup> degree burn [%] § | 22 (68.8) |  |
| inhalation trauma [%] § | 7 (21.9) |  |

\* indicated are mean (median) ± SD [interquartile range]
§ indicated are n (SD)

Sera were obtained daily within the first week after admission, and weekly up to day 28 after admission. For controls, samples were obtained in the same manner. Whole blood was incubated for thirty minutes at ambient temperature and centrifuged at 2850 relative centrifugal force for 17 minutes. Sera were separated and cryopreserved below -70°C.

Intensive care management including fluid resuscitation was performed according to institutional guidelines and was not affected by this study. Inhalation trauma was assessed on primary survey and deemed present in cases of visible signs of thermal injury within the upper airway. Mortality was defined as all-cause in-hospital mortality.

### Analyte quantifications

Serum MPO, CitH3, NE, C3a, and lactadherin (milk fat globulin protein E8, MFG-E8) were quantified by enzyme-linked immunosorbent assay (ELISA) using commercially available kits as recommended by the manufacturers (human MPO, human neutrophil elastase, human MFG-E8 Immunoassay Quantikine, all R&D Systems, Bio-Techne, Minneapolis, MN, USA; human CitH3, clone 11D3, Cayman Chemical, Ann Arbor, MI, USA; human complement C3a, Invitrogen, Thermo Fisher Scientific, Waltham, MA, USA). Colorimetric measurements were performed using Tecan F50 infinite microplate reader (Tecan Group, Männedorf, Switzerland) with Magellan software (version 7.2, Tecan). Analyte concentrations were determined by external standard curves.

### Laboratory measurements of clinical parameters

Complete blood counts were determined during routine clinical tests and reference values were adopted from in-house reference ranges. Sequential organ failure assessment (SOFA) score (34), acute physiology and chronic health evaluation II (APACHE II) score (35), and abbreviated burn severity index (ABSI) (36) were assessed on the day of admission. Median values of severity scores were used to assign values to low and high groups. As the SOFA score of our study population displayed a median of 6.5, we chose ≤ 6 and ≥ 7 as cut-off values.

### Statistical analyses

Data were statistically evaluated and visualized using SPSS Statistics (version 25, IBM, Armonk, NY, USA) and GraphPad Prism (version 5.01, GraphPad Software Inc., La Jolla, CA, USA). Continuous variables were compared by the Mann-Whitney test. One-way ANOVA and multiple comparison *post hoc* tests with Sidak’s or Dunnett’s correction were calculated. For correlation analysis, Pearson’s correlation coefficients were calculated. Data are presented as arithmetic means ± standard error of the mean.

## Results

### Neutrophil-derived factors are systemically elevated following burn injury

To study the role of burn injury-induced NETosis, we tracked the neutrophil-derived factors MPO, CitH3, NE, and C3a in sera of burn victims up to 4 weeks post trauma. We found that NETosis-associated factors were significantly increased compared to healthy controls (Figure 1). MPO was remarkably increased in the early days post burn injury (Figure 1A). While CitH3 did not differ from controls on admission day and day 1, serum CitH3 concentrations showed a delayed increase, peaking on day 4 post admission (Figure 1B), indicating a potential ‘second burn’ hit. Intriguingly, NE levels were elevated on admission day and in the early stress response to burn trauma before approximating levels detected in healthy controls (Figure 1C). Serum C3a was strongly elevated in burn victims compared to controls (Figure 1D). To test a potential connection between systemic C3a levels and liver parameters in burn victims, we furthermore assessed gamma-glutamyltransferase (γ-GT) levels. γ-GT started increasing 2 weeks post injury, but did not correlate with C3a (Supplemental Figure S1). Since lactadherin promoted survival of septic rats (37) and lactadherin prevented coagulopathy following traumatic brain injury (38), we addressed the question whether lactadherin might also be involved in the burn injury-induced immune response. When assessing serum lactadherin levels, we observed no difference between burn victims and healthy controls (Supplemental Figure S2). We furthermore aimed to investigate the relationship between neutrophil-derived factors and found a positive correlation between MPO and NE, but not between CitH3 and NE or between CitH3 and MPO (Supplemental Figure S3). These data suggest a role for NETosis in the systemic post burn injury immune response.

**Figure 1.**
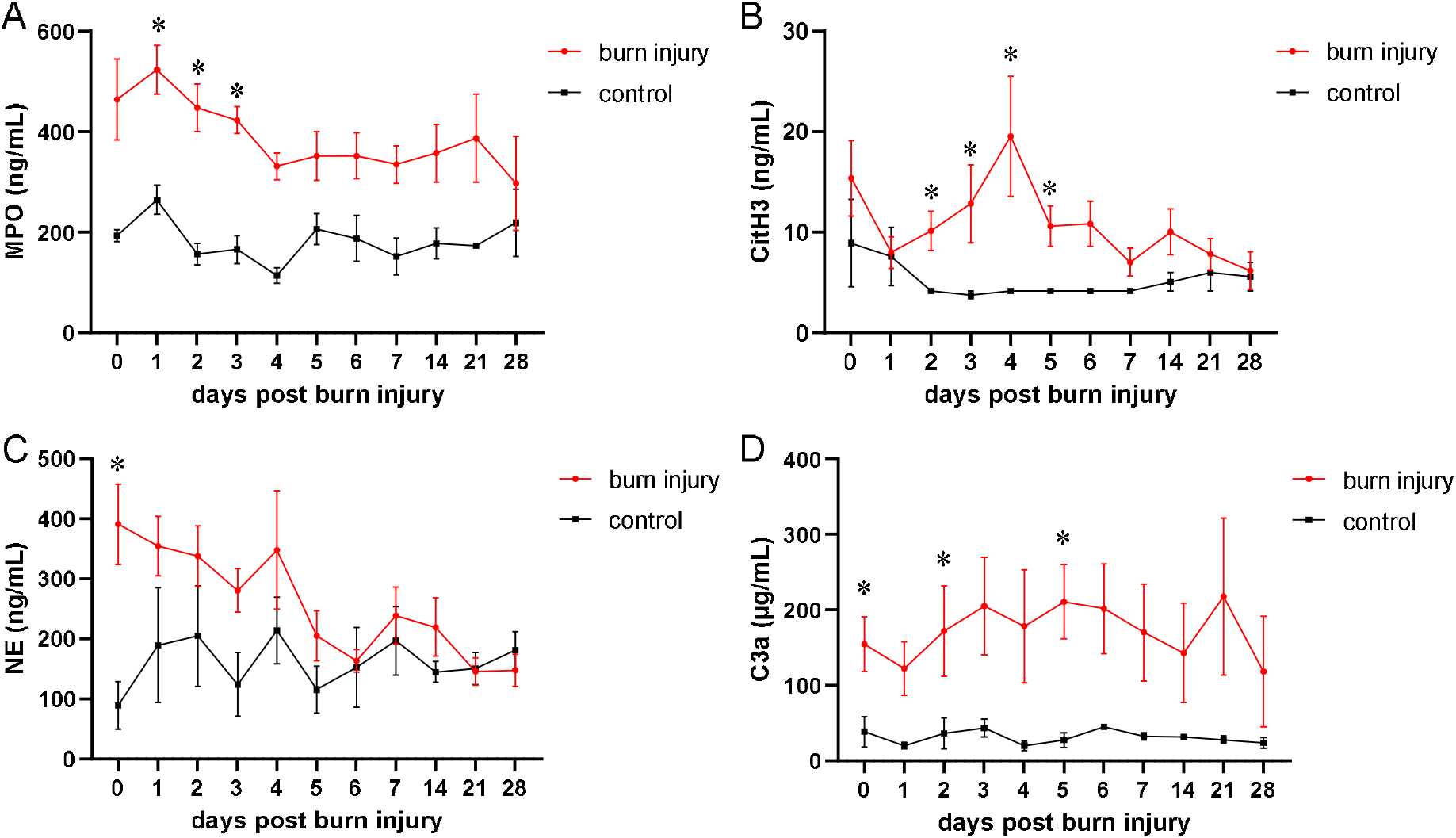
Elevated neutrophil-derived factors and C3a in burn sera. Systemic levels of (A) MPO, (B) CitH3, (C) NE, and (D) C3a were quantified in sera of burn victims and healthy controls up to 4 weeks post injury. Data were compared by mixed effects analysis and Sidak’s multiple comparisons *post hoc* test. Data are presented as means ± s.e.m. Asterisks indicate *p* values <.05.

### Burn injury induced leukocytosis and relative lymphopenia

Next, we assessed immune cell status in our patient cohort. We found that absolute leukocyte and neutrophil numbers were strongly increased in burn victims (Figures 2A, C, & D). In detail, cell numbers were elevated in the early days post injury, decreased within the first week before incrementing again. While platelet counts of burn victims were below those of healthy controls within the first week post injury, we observed a remarkable increase in platelet numbers 3 weeks post trauma (Figure 2B). These data suggest a delayed, immunological ‘second hit’ and burn injury progression even weeks after primary injury. Absolute lymphocyte counts showed no difference between controls and burn victims, while relative lymphocyte levels were strongly decreased in the first 7 days after injury (Figures 2E & F). Together, these data indicate an intricate systemic effect of burn injury on immune cell status.

**Figure 2.**
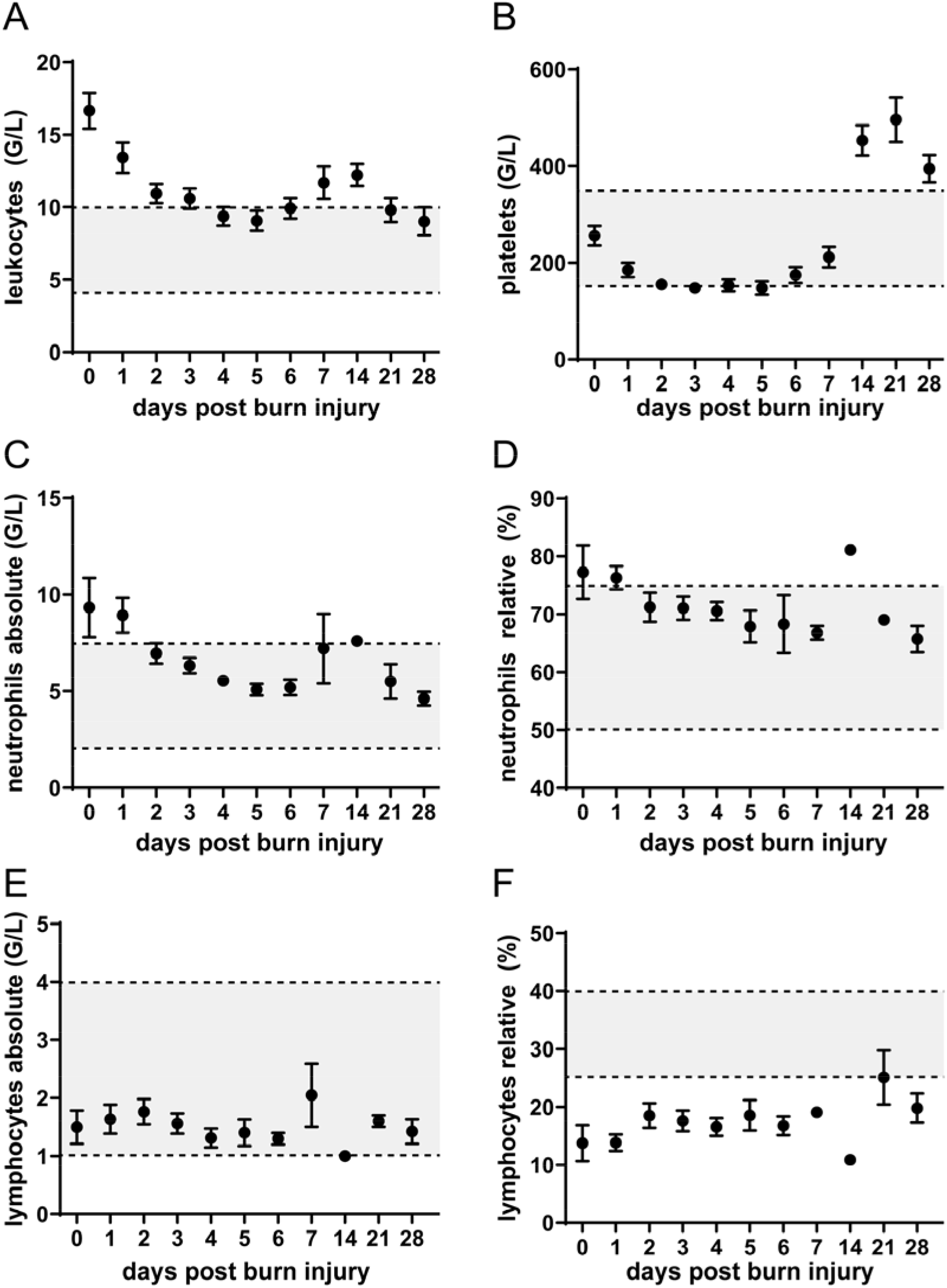
Immune cell counts up to 4 weeks post injury. Patient data in relation to established in-hospital reference standards. Data are presented as means ± s.e.m.

### 3^rd^ degree burns are associated with increased CitH3 and NE

We aimed to test whether our set of serum markers might be associated with mortality, inhalation trauma, or 3^rd^ degree burn injury. Our battery of neutrophil-derived factors did not predict mortality, and C3a was even strongly reduced in deceased burn victims (Supplemental Figure S4). We furthermore compared burn victims with inhalation trauma to those without respiratory damage and observed no effect of lung injury on serum levels of neutrophil-derived factors (Supplemental Figure S5). Similarly, serum MPO levels displayed no difference when comparing 3^rd^ degree burn victims with patients suffering from 1^st^ and 2^nd^ degree burns (Figure 3A). By comparison, we observed elevated serum CitH3 and NE levels on days 3 and 4 of 3^rd^ degree burns compared to first and second degree burns (Figures 3B & C). Of note, the delayed increase of serum CitH3 3 and 4 days post injury were exclusively detected in 3^rd^ degree burns. Surprisingly, C3a levels were strongly reduced in higher degree burns on day 3 (Figure 3D). These findings suggest that the degree of burn injury determines the extent of neutrophil activation and NETosis.

**Figure 3.**
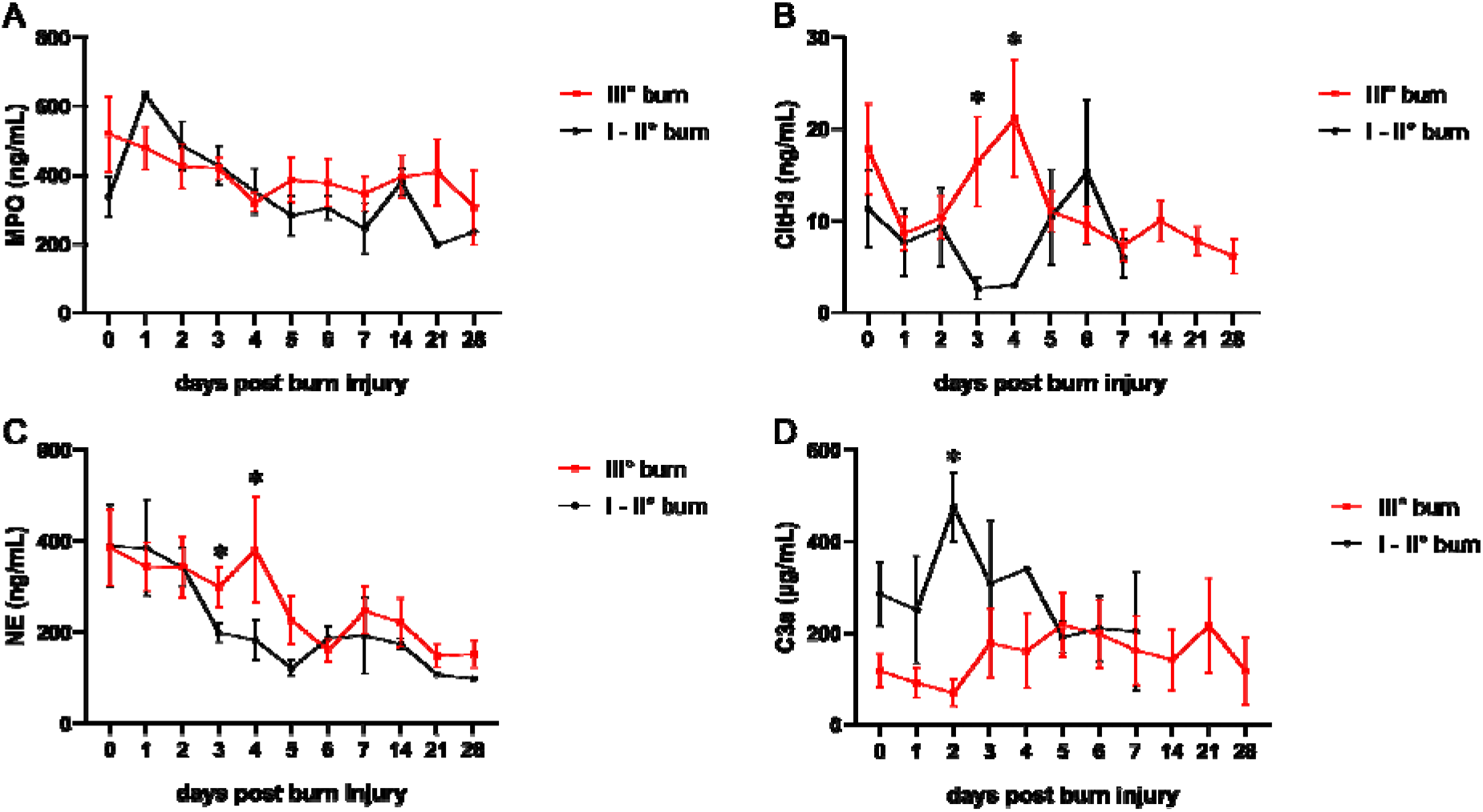
Increased NETosis in higher degree burns. Serum concentrations of (A) MPO, (B) CitH3, (C) NE, and (D) C3a in 3^rd^ degree burn victims compared to lower degree burns. Data are presented as means ± s.e.m. ^§^ indicates *p* values <.05.

### CitH3 is elevated in patients with higher severity scores

We sought to determine whether neutrophil-derived factors were associated with clinical severity scoring systems and dichotomized SOFA, APACHE II, and ABSI scores into low and high values. We observed increased MPO and CitH3 levels in patients with APACHE II scores above 18 (Supplemental Figure S6) and NE was significantly higher in SOFA scores ≥ 7 (Supplemental Figure S7). Intriguingly, CitH3 and NE levels were elevated in patients with ABSI scores above 9, while MPO and C3a showed no difference (Figure 4). These data suggest that specific NETosis-associated factors are elevated in more severe burns.

**Figure 4.**
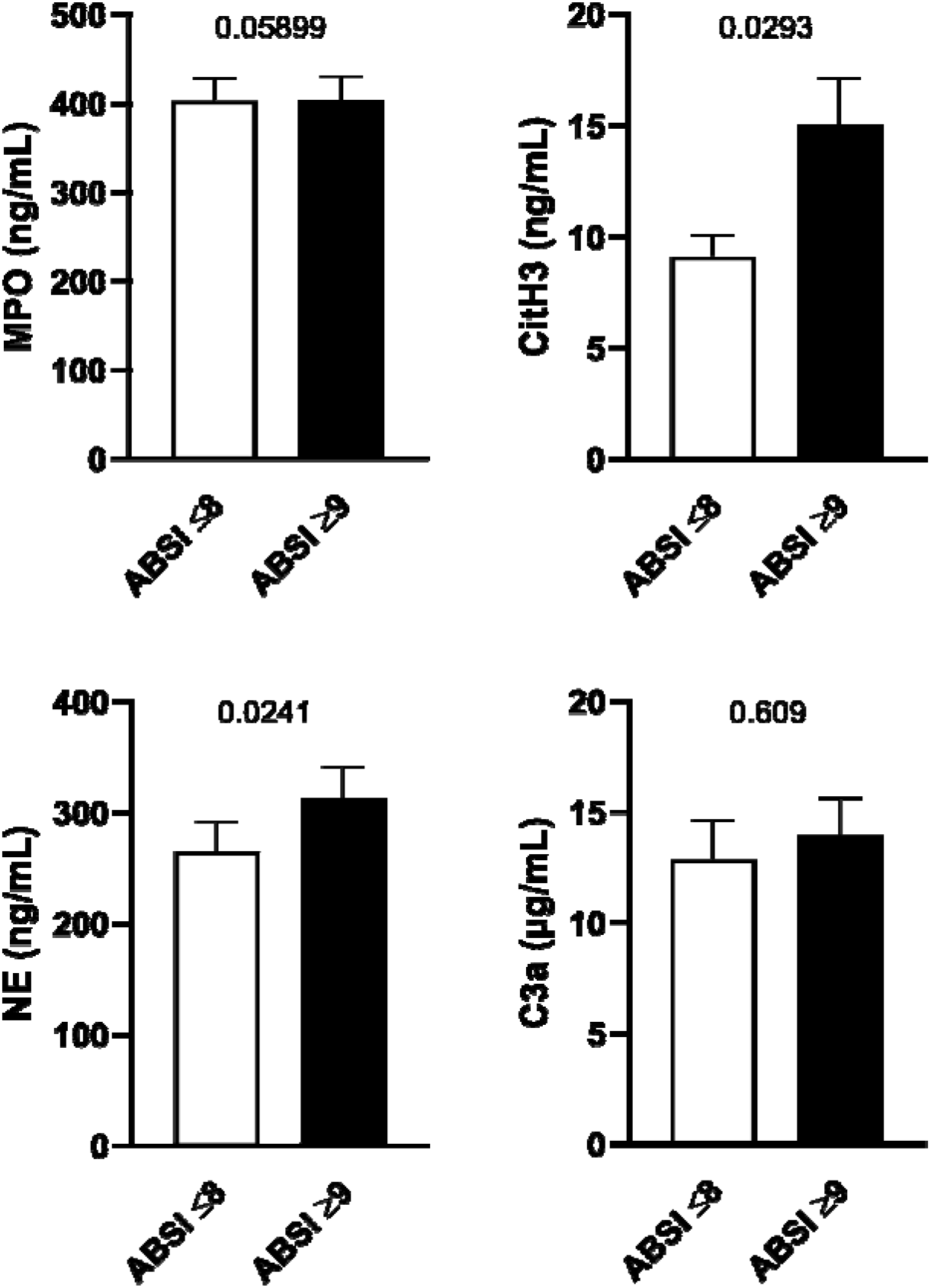
Serum concentrations of neutrophil-derived factors in low and high ABSI scores. ABSI scores were determined on admission day and categorized into low (≤ 8) and high (≥ 9) values. Serum concentrations of MPO, CitH3, NE, and C3a of the first 7 days post admission were compared. Data were statistically evaluated by Mann-Whitney test.

### Burn injury immune cell signature is independent of severity

Finally, we investigated whether the burn injury-induced alterations in immune cell levels were associated with severity of burn injury. Therefore, cell counts were compared between 3^rd^ degree and lower degree burn victims and furthermore between patients with higher and lower clinical severity scores. A trend towards increased leukocyte and neutrophil counts in more severe cases was found on admission day, while no difference in platelet and relative neutrophil counts was observed (Supplemental Figures S8 - S11). Absolute and relative lymphopenia was more pronounced in the early days following burn injury when comparing low versus high SOFA scores (Supplemental Figure S10), indicating that immune cell counts are time- and score-dependent.

## Discussion

Burn injuries trigger a complex immune response characterized by a unique cytokine secretion pattern and immune cell activation. Exaggerated burn-induced immune activation can lead to tissue damage and organ dysfunction, while immunosuppression can predispose to infectious diseases. Activation of neutrophils and secretion of neutrophil-derived factors add another piece to the complex picture of immunological reactions in response to burn injuries. We were the first to track systemic levels of neutrophil-derived immunomodulators up to 4 weeks post trauma and were able to demonstrate that the NETosis surrogate markers MPO, CitH3, NE, and C3a were increased in burn victims compared to healthy controls. Several studies have already suggested a role for neutrophils in the immune response elicited by burn injuries. Circulating DNA in plasma has been identified as a prognostic marker for mortality and an early biomarker of sepsis in burn-injured patients (12, 28), though free DNA does not exclusively originate from NETs (39) and is therefore considered a rather indirect evidence of neutrophil involvement. Further studies investigated more neutrophil-specific factors. Bacteriostatic effects of topically administered MPO were demonstrated in *Staphylococcus aureus*-infected burn wounds (40) and increased MPO activity was observed following burn injury in rats (41). Intriguingly, burn injury reduced NETosis, indicated by diminished CitH3 levels, *via* pulmonary immunosuppression (31). A role for NE in the proteolytic degradation of fibronectin in burn wound fluids (42) and in burn-blast-induced lung injury in rats (43) has been reported and inhibiting NE activity exerted cytoprotective effects (43).These studies investigated local levels of MPO, CitH3, and NE in burn wounds, burn fluids, or lung tissues. Systemic concentrations of these factors have not been comprehensively described to date. Our data with a 4 week follow up time post burn injury therefore add valuable information to the role of NETosis in the burn injury-induced stress response.

We detected increased serum concentrations of complement C3a in burn-injured patients compared to controls. Elevated C3a levels have already been reported previously in plasma of burn victims (44) and in subdermal tissues of second-degree thermal wounds (45). Interestingly, expression of complement receptors on neutrophils was increased following burn injury (46) and it was previously demonstrated that neutrophilic elastase promotes complement amplification (47). Hence, an intricate crosstalk between the complement system and neutrophils occurs in the immune response to burn injury. Though intracellular stores of C3a have been found in neutrophils (20), numerous other cell types and tissues, such as the liver (48), are conceivable to contribute to serum C3a levels post burn injury. Our C3a levels did not correlate with γ-GT, indicating that complement activation is not associated with liver disease in our settings. Delineating the exact cellular origin of systemic C3a in burn victims will be subject of future studies.

We observed remarkably increased absolute leukocyte counts in burn victims compared to healthy controls, while absolute lymphocyte counts remained largely unaltered. In addition, absolute neutrophil levels were strongly increased post burn injury. These data are in line with data reported by Mulder *et al*. (49) and it is tempting to speculate that the elevated neutrophil amounts detected in our patient cohort might serve as a potential cellular source for MPO, CitH3, NE, and C3a. We furthermore observed that the increase of absolute leukocyte and neutrophil counts was even more pronounced in more severe cases. Intriguingly, relative lymphocyte levels were remarkably decreased in patients with burn injury and lower in patients with higher SOFA score. Lymphopenia has already been reported in septic and burn victims, which might contribute to post-injury immunosuppression, morbidity, and mortality (50-52). Platelet levels were decreased in the early stress response and re-elevated starting 14 days after burn injury. These dynamics are in accordance with previous reports, where a sustained pro-coagulant state of burn patients was described (53). Together, these data connect the systemic factors with clinical blood parameters and contribute to a better understanding of the cellular and molecular processes in the concerted burn injury response.

Microvascular obstruction was reported as the leading cause of burn injury progression and red blood cell aggregates, microthrombi, and ischemia have been described to promote secondary tissue damage (16). NETs serve as a scaffold for platelets, induce platelet aggregation and coagulation (23, 24) and have furthermore been shown to exert pro-coagulant activities in sepsis (25). Moreover, neutrophil activation and CitH3 were induced by myocardial ischemia/reperfusion injury and showed pro-thrombotic and cytotoxic features, while eliminating NETs by DNase I treatment exerted cardio-protective effects (26). In addition, elevated CitH3 levels were reported in various inflammatory conditions with a role of microvascular thrombosis (27, 54). Though lactadherin levels remain unaltered, we observed systemic CitH3 elevation 2 – 5 days post burn injury and detected abnormal immune cell counts even several weeks after burn injury. These findings corroborate the concept of burn injury progression following initial damage. We hypothesize that injury-induced elevation of neutrophil counts and concomitant neutrophil activation contribute to burn injury progression. NETosis and systemic neutrophil-derived factors are induced in the course of a delayed immunological response, provoke microvascular obstruction, and thereby promote secondary tissue damage.

Our data revealed that 3^rd^ degree burns showed higher CitH3 and NE levels and these results are in line with the clinical severity score ABSI, where cases with higher scoring displayed increased CitH3 and NE. These data suggest that the severity of burn injury dictates the level of neutrophil activation. Intriguingly, patients succumbing to burn trauma did not display elevated neutrophil-derived immunomodulators. While we found elevated CitH3 and NE levels in patients with higher degree and severity burns, future studies with larger samples sizes will be required to determine whether elevated NETosis-associated factors might serve as a prognostic factor for mortality following severe burn injury. Moreover, no difference between burn victims suffering from inhalation trauma compared to burn-injured patients without inhalation trauma was observed. Neutrophilia in the bronchoalveolar space upon inhalation trauma has been described and attenuating neutrophil recruitment improved histopathology scores and bacterial clearance (55). Though NETosis markers were not related to inhalation trauma in our patient collective, a role for burn-associated lung injury in neutrophil activation cannot be entirely ruled out (56).

In spite of our best efforts, this study has certain limitations. Though 32 burn victims were initially included, the number of available specimen declined over time as patients either succumbed to their injury or were discharged from the hospital. Future studies with higher sample numbers will be required to fully elucidate the role of neutrophil-derived factors in the burn injury response. Our set of neutrophil-derived factors serves as a surrogate machinery to assess NETosis. Further investigations are necessary to provide more direct evidence of systemic NETosis in sera of burn victims.

Clinical management of burn victims involves fluid resuscitation, burn wound coverage, supportive care, and rehabilitation. Targeting neutrophil function in post burn injury might serve as an alternative treatment option in severe cases with exacerbated neutrophil activation. Determining the full therapeutic potential of NETosis inhibition following burn trauma will merit future investigations.

## Data Availability

Raw data are available from the corresponding authors upon request.

## Data Availability Statement

Raw data are available from the corresponding authors upon request.

## Author contributions

Idea & Conceptualization: HJA and ML; Data curation, ML, MTL, and TH; Formal analysis, ML and MTL; Funding acquisition, HJA; Investigation, ML, MTL, and TH; Project administration, HJA and TH; Resources, HJA and TH; Supervision, HJA and TH; Writing – original draft, ML, HJA, and TH; Writing – review & editing, MTL, DC, MD, KK, DB, AG, BM, CR, SH, and MM.

## Acknowledgements

We thank the Dr. HP Haselsteiner and the CRISCAR Familienstiftung for their belief in this private public partnership to augment basic and translational clinical research.

