## Supplemental Information for "Severity of thermal burn injury is associated with systemic neutrophil activation"

**Supplemental Figure S1**

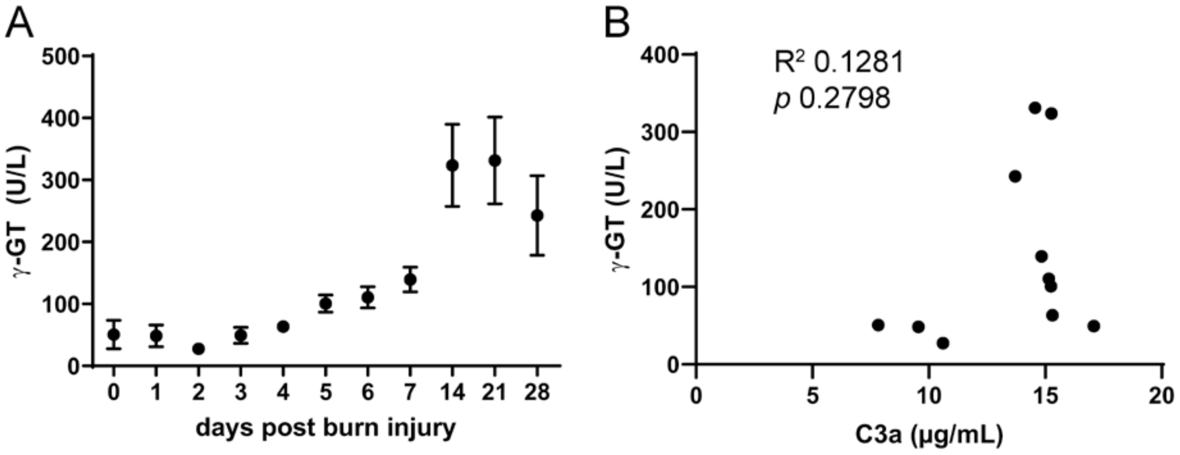


**Supplemental Figure S1. γ-GT levels following burn injury.** (A) Serum γ-GT levels in burn victims up to 4 weeks post injury. (B) Correlation between serum C3a and γ-GT concentrations over 28 days post burn trauma.

**Supplemental Figure S2**

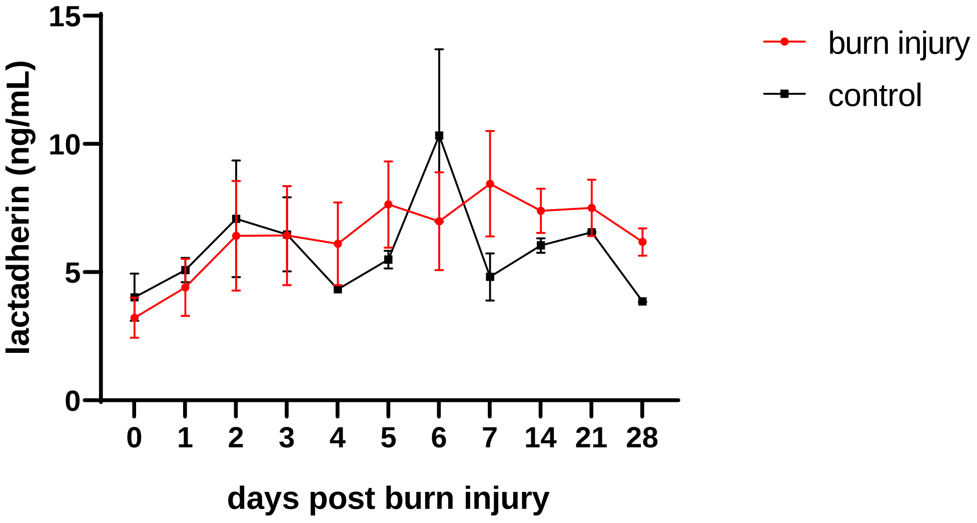


**Supplemental Figure S2. Lactadherin levels following burn injury.** Lactadherin concentrations in sera of burn victims and healthy controls over 4 weeks.

**Supplemental Figure S3**

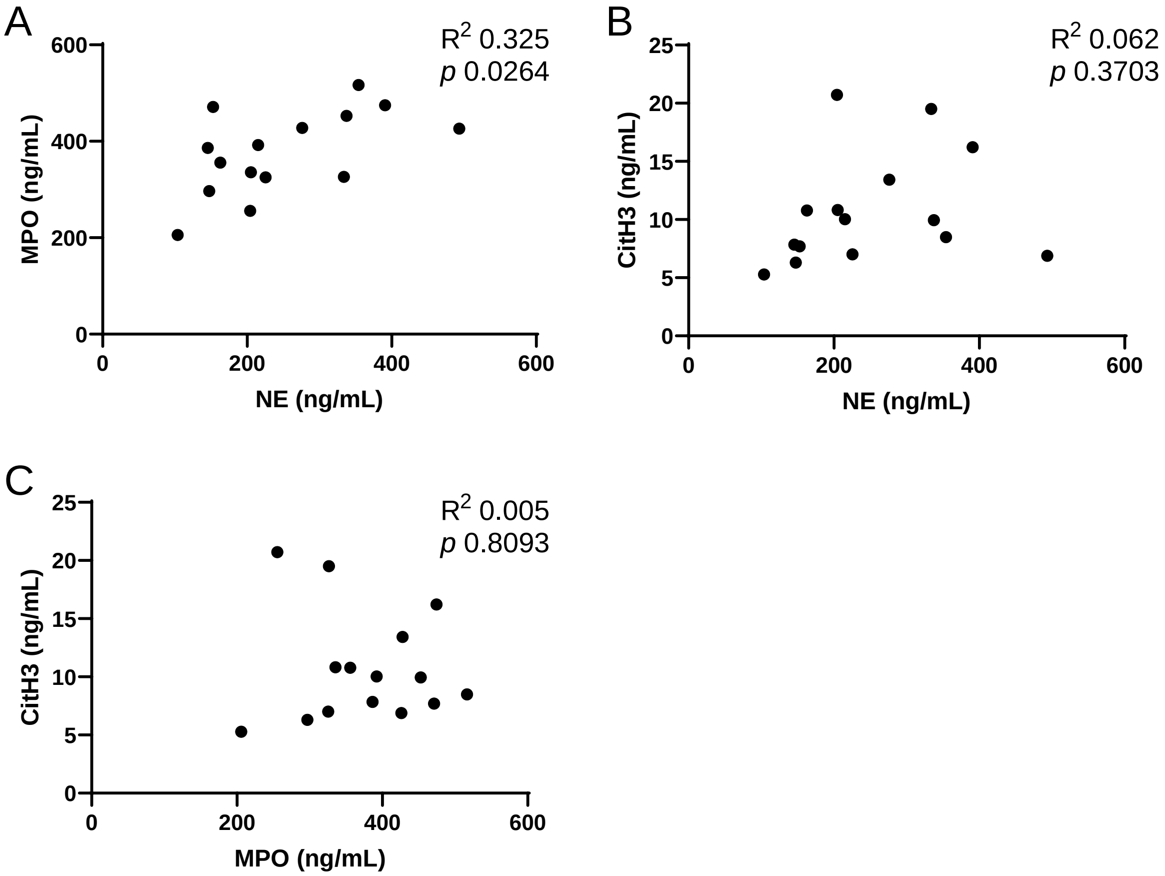


**Supplemental Figure S3. Correlations between serum levels of NETosis markers in burn victims.** Correlation between (A) MPO and NE, (B) CitH3 and NE, and (C) CitH3 and MPO on admission day.

### Supplemental Figure S4

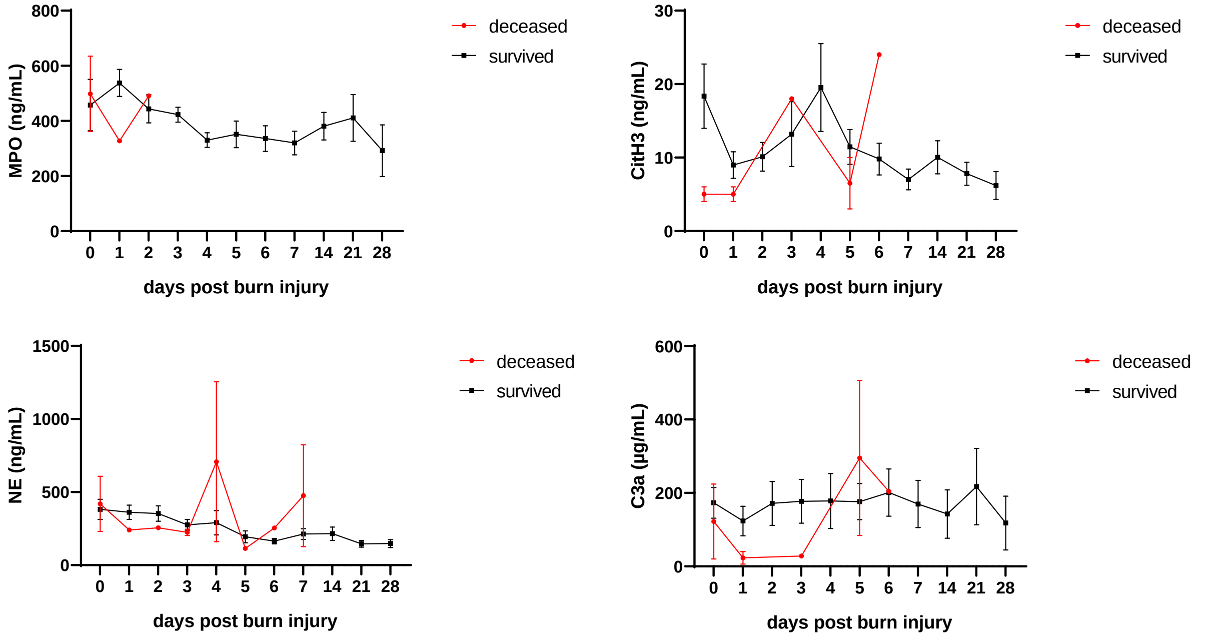


**Supplemental Figure S4. NETosis levels in patients succumbing to burn injury.** Serum concentrations of neutrophil-derived factors were compared between survivors and deceased patients.

### Supplemental Figure S5

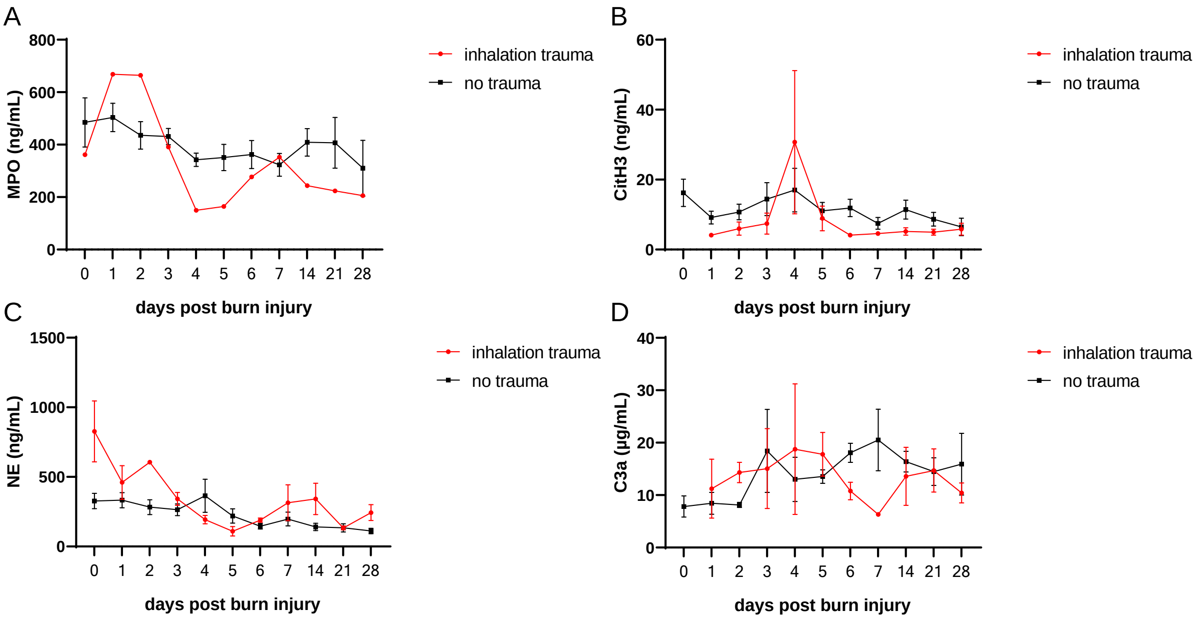


**Supplemental Figure S5. Effect of inhalation trauma on serum NETosis levels.** Serum levels of neutrophil-derived factors were compared between burn victims with inhalation trauma and without inhalation trauma. Serum levels of (A) MPO, (B) CitH3, (C) NE, and (D) C3a were assessed up to 4 weeks post burn injury.

**Supplemental Figure S6**

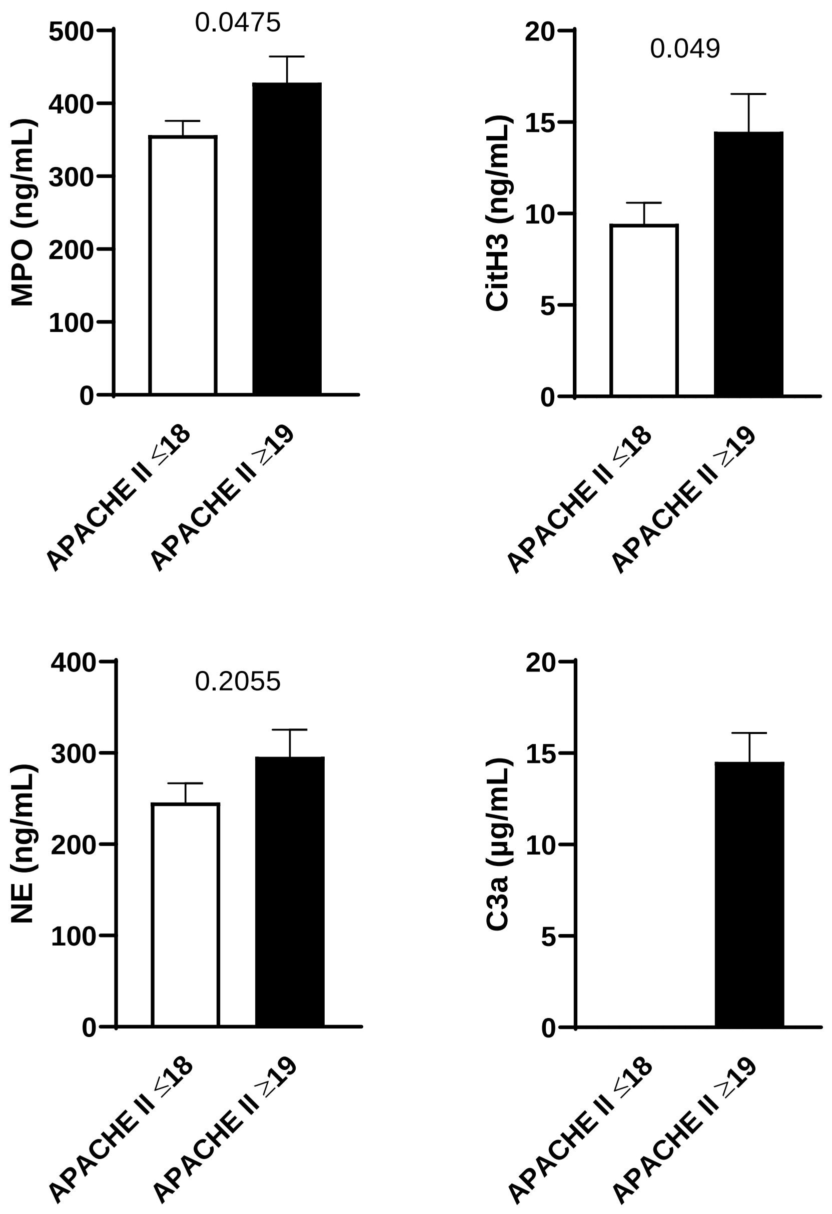


**Supplemental Figure S6. Serum concentrations of neutrophil-derived factors in low and high APACHE II scores.** Scores were determined on admission day and categorized into low (≤18) and high (≥19) values. Serum concentrations of MPO, CitH3, NE, and C3a of the first 7 days post admission were compared. Data were compared by Mann-Whitney test.

**Supplemental Figure S7**

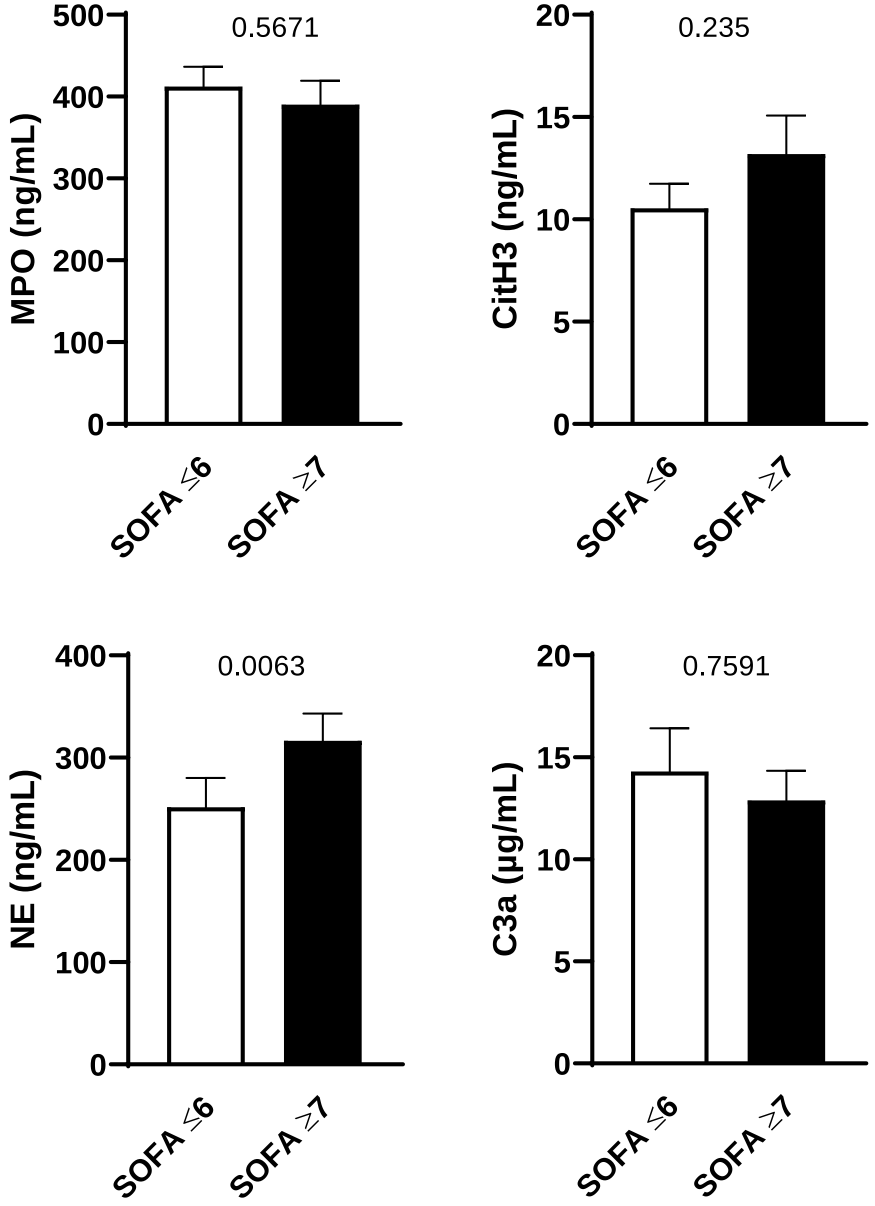


**Supplemental Figure S7. Serum concentrations of neutrophil-derived factors in low and high SOFA scores.** Scores were determined on admission day and categorized into low (≤6) and high (≥7) values. Serum concentrations of MPO, CitH3, NE, and C3a of the first 7 days post admission were compared. Data were compared by Mann-Whitney test.

### Supplemental Figure S8

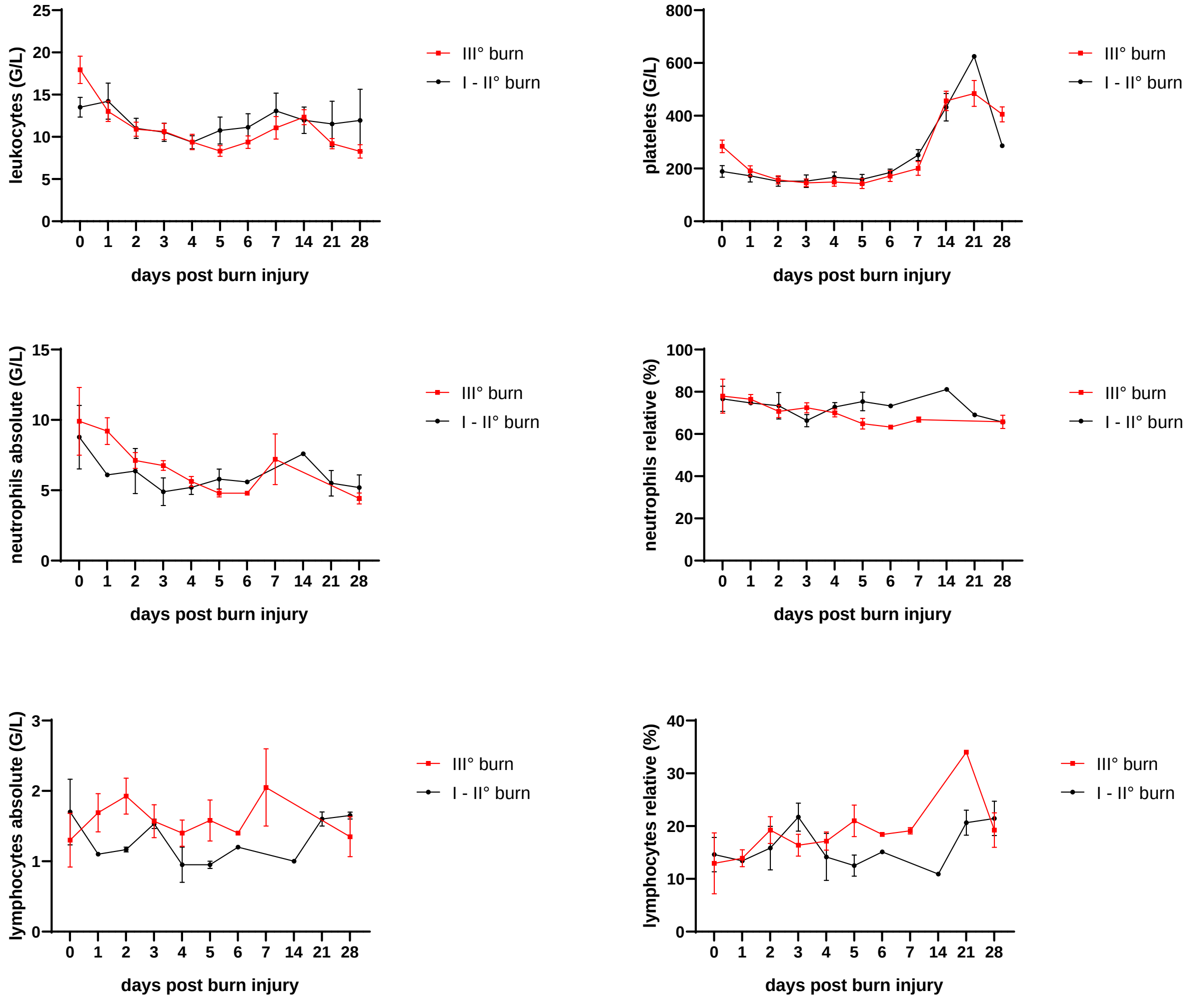


**Supplemental Figure S8. Immune cell counts of patients with higher and lower degree burns.** Data of 3rd degree burns and lower degree burns were compared by Mann-Whitney test.

### Supplemental Figure S9

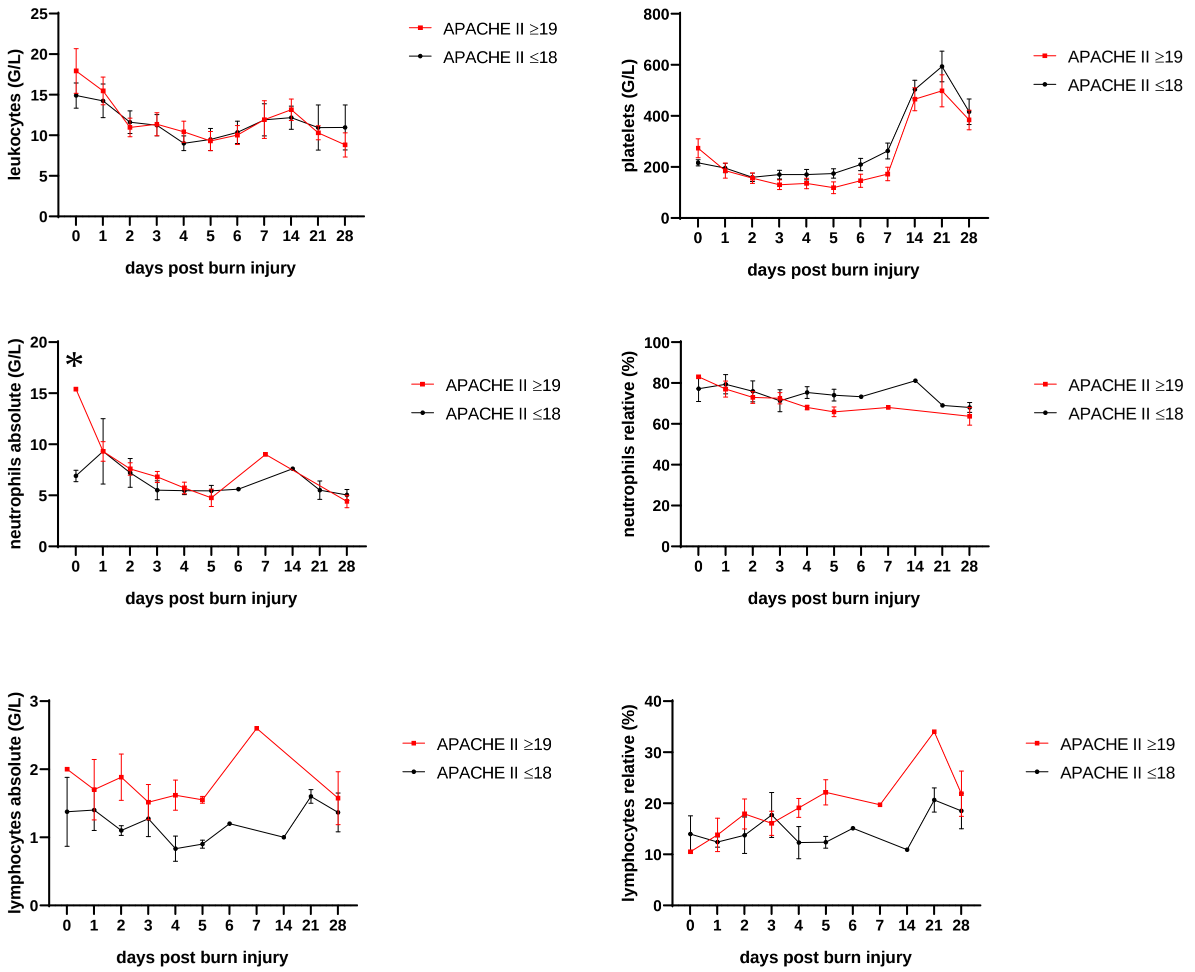


**Supplemental Figure S9. Immune cell counts of patients with high and low APACHE II scores.** Scores were determined on admission day and categorized into low (≤ 18) and high (≥ 19) values. Data were compared by Mann-Whitney test.

### Supplemental Figure S10

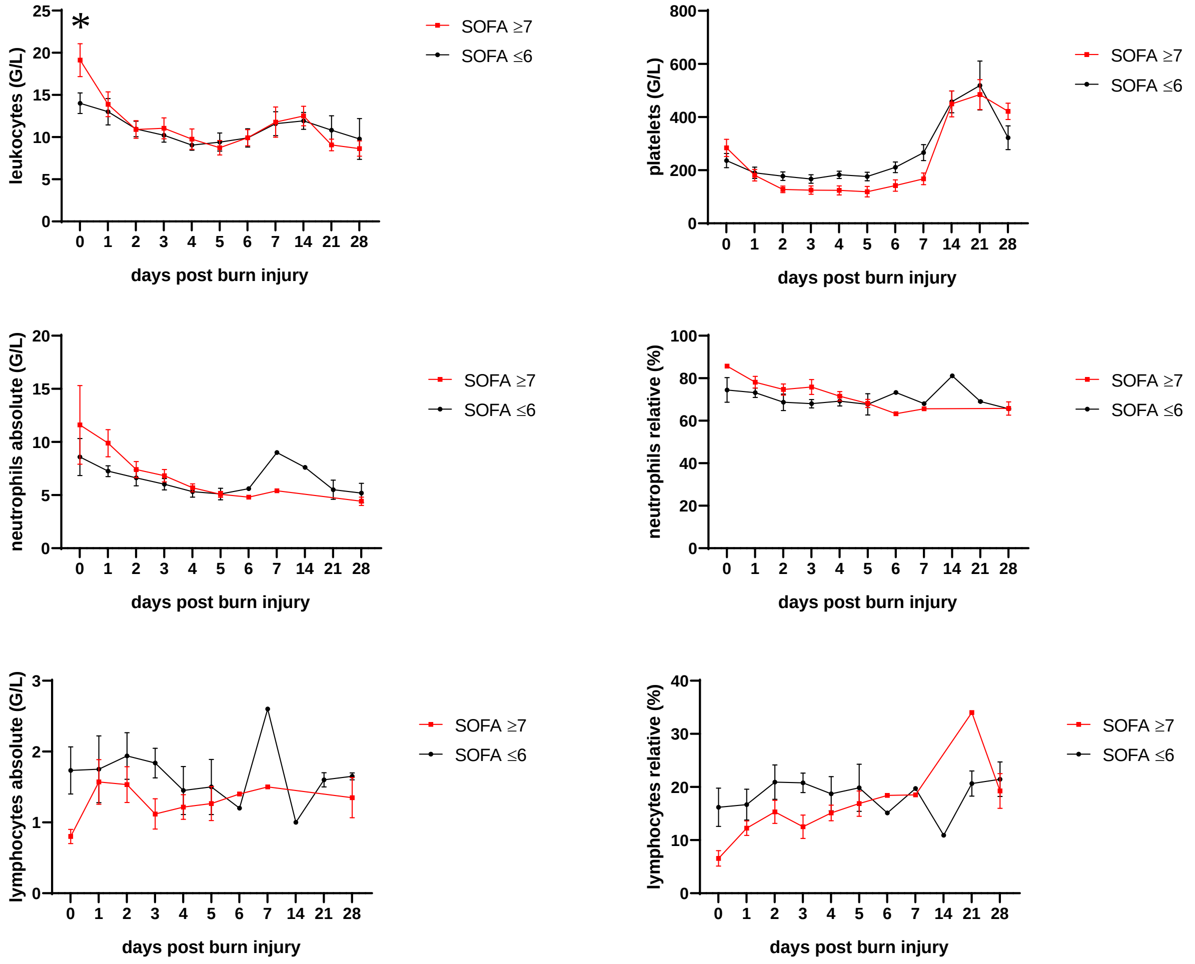


**Supplemental Figure S10. Immune cell counts of patients with high and low SOFA scores.** Scores were determined on admission day and categorized into low (≤ 6) and high (≥ 7) values. Data were compared by Mann-Whitney test.

### Supplemental Figure S11

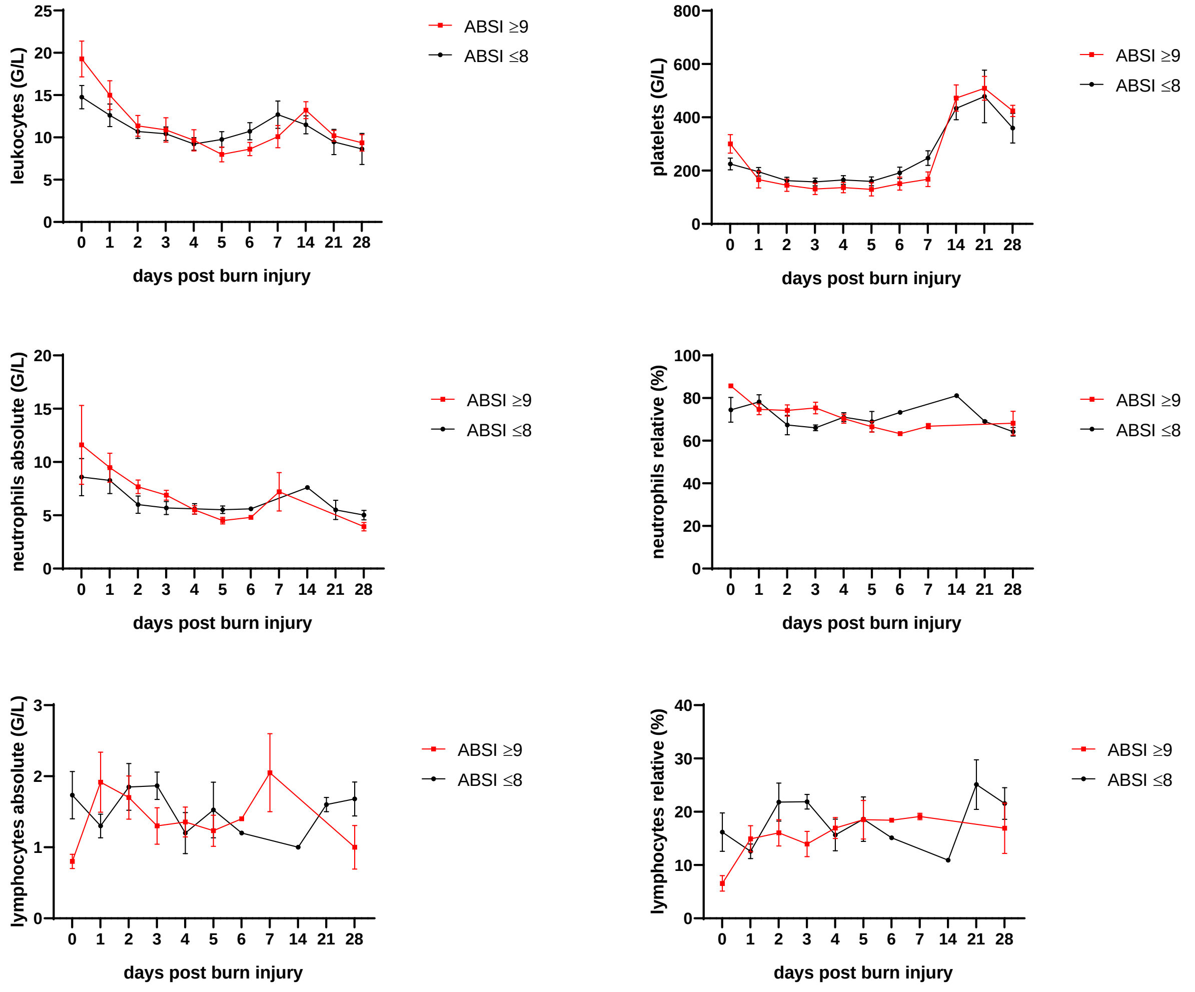


**Supplemental Figure S11. Immune cell counts of patients with high and low ABSI scores.** Scores were determined on admission day and categorized into low (≤ 8) and high (≥ 9) values. Data were compared by Mann-Whitney test.
